# *TMPRSS2* variants and their susceptibility to COVID-19: focus in East Asian and European populations

**DOI:** 10.1101/2020.06.09.20126680

**Authors:** Ney Pereira Carneiro dos Santos, André Salim Khayat, Juliana Carla Gomes Rodrigues, Pablo Pinto, Gilderlanio Santana de Araújo, Lucas Favacho Pastana, Jéssyca Amanda Gomes Medeiros, Marianne Rodrigues Fernandes, Arthur Ribeiro-dos-Santos, Bruna Claudia Meireles Khayat, Fabiano Cordeiro Moreira, André Maurício Ribeiro-dos-Santos, Paula Baraúna de Assumpção, Ândrea Ribeiro-dos-Santos, Paulo Pimentel de Assumpção, Sidney Santos

## Abstract

The manifestation of the COVID-19 varies from absence of symptoms to Severe Acute Respiratory Syndrome. The epidemiological data indicate that infection and mortality rates are greater in European populations in comparison with eastern Asians. To test if epidemiological patterns may be partly determined by human genetic variation, we investigated, by exomic and databank analyses, the variability found in the *TMPRSS2* gene in populations from different continents, since this gene is fundamental to virus access into human cells. The functional variants revealed low diversity. The analyses of the variation in the modifiers of gene expression indicate that the European populations may have much higher levels of pulmonary expression of the *TMPRSS2* gene and would be more vulnerable to infection by SARS-CoV-2. By contrast, the pulmonary expression of the *TMPRSS2* may be reduced in the populations from East Asia, which implies that they are less susceptible to the virus infection and, these genetic features might also favor their better outcomes. The presented data, if confirmed, indicates a potential genetic contribution of *TMPRSS2* to individual susceptibility to viral infection, and might also influence COVID-19 outcome.

## 1. Introduction

Coronavirus disease 2019 (COVID-19) is a severe disorder caused by a single-stranded RNA beta-coronavirus (SARS-CoV-2) whose genome is 90% similar to that of the SARS-CoV virus, responsible for the 2002 epidemic, and 50% similar to MERS-CoV, which caused the 2012 epidemic [1, 2]. The first cases of COVID-19 were recorded in the Chinese city of Wuhan, in December 2019. Given the high potential for the transmission of the disease, the World Health Organization (WHO) declared the outbreak a pandemic on March 11th 2020 [3, 4]. At the present time, more than six million cases have been registered, worldwide, with more than 370,000 deaths [5, 6, 7].

Patients infected with COVID-19 may present an ample spectrum of manifestations, ranging from an absence of any major clinical symptoms, as seen in the majority of cases, to Severe Acute Respiratory Syndrome, which is often fatal [8]. This diversity of clinical outcomes is found not only between individuals, but also between populations within the same country, among countries on the same continent, and even among continents [9].

The mean number of cases per 100,000 inhabitants may vary considerably among populations, with rates over 270 infections per 100,000 inhabitants in Europe and 180 in Asia. In Europe, these rates also vary considerably among countries, from >500 infections per 100,000 inhabitants in Spain and Belgium, 340 in Italy to 190 in Germany. The discrepancies in the death rates are even greater, with rates over 20 deaths per 100,000 inhabitants in Europe, seven in America, and below two in Asia. These numbers also vary significantly among European countries, such as Germany (nearly 10 deaths per 100,000 inhabitants), Spain, Italy and United Kingdom (almost 60) and Belgium (over 80) [9].

A *priori*, this variation in infection and death rates may be related primarily to differences in factors such as the isolation policies in the different countries or populations, access to medical care, the age structure of the population, and the reliability and scope of the epidemiological data [10]. However, these factors do not appear to account fully for the discrepancies observed among populations, and important lacunas exist, which demand the attention of the scientific community, for the proposal and testing of hypotheses that consolidate the understanding of the observed pattern. Although the numbers of cases and deaths may be under-reported for some populations, the variation observed among countries known to have rigorous standards for the compilation and presentation of epidemiological data indicates that human genetic variability should be considered a likely candidate to justify some of the discrepancies in the data on both infection and death rates.

Two genes in particular have received special attention from researchers, due to their role in the invasion of human cells by the virus. One is the *ACE2* gene, which codifies a cell receptor that allows the viral S protein (spike) to bind to the cell, and the other is the *TMPRSS2* gene which codifies a serine protease that interacts with the viral S protein, permitting the fusion of the cell and viral membranes [11]. The role of these genes has already been recognized in the infections by SARS-CoV and MERS-CoV [12, 13, 14] and, more recently [15, 16], SARS-CoV-2 has been found to use the same mechanisms (*ACE2* and *TMPRSS2*) to bind to human cells. In addition to exploiting similar access routes, the infection by SARS-CoV, MERS-CoV and SARS-CoV-2 also shares the tropism through the upper respiratory air passages, which is fundamental to the pulmonary pathogenesis, a basic characteristic of these infections [17, 18, 19]. Given this, the tissue of the upper respiratory passages, which express *ACE2* and *TMPRSS2* together, would have a greater probability of being a target for the access of these viruses to the human organism.

A number of different research groups have investigated the potential associations between structural genetic markers of the *ACE2* gene and both infection rates and the clinical manifestations of COVID-19 [11, 20, 21, 22]. Our research team recently presented a detailed review of the variation in this gene in the different human populations around the world [23], and found important differences among populations genetic polymorphisms, with potentially serious implications for COVID-19 outcomes.

The *TMPRSS2* gene is amply expressed in epithelial tissue that lines upper air passages, bronchi, and lungs, as well as in other types of tissue [24]. This gene is expressed in combination with *ACE2* in the type II pneumocytes, which are known to be viral targets [25, 11], in bronchial transient secretory cells [18], and in the cells of the olfactory epithelium [19].

A high expression of the *TMPRSS2* gene may be associated directly with the COVID-19 infection pattern. Patients with prostate cancer are known to have an over-expression of *TMPRSS2* in both the prostate and the lungs [26]. A recent study [27] showed that Italian prostate cancer patients being treated with Androgen Deprivation Therapy (ADT) are partially protected from infection by SARS-CoV-2, probably due to the reduction in the expression of *TMPRSS2* by this medication.

Given the importance of the *TMPRSS2* gene for the process of infection by SARS-CoV-2, the present study investigated genetic markers potentially responsible for alterations in the function or expression of the gene in the lungs, and that have a high degree of variability among the populations, which may be linked to the differences observed in the susceptibility to infection and the severity of the outcome in the different populations, around the world. The potential identification of markers for the risk of infection, development of specific symptoms and diverse outcomes would be extremely valuable to public health authorities and medical treatment facilities throughout the world, and should represent a fundamentally important tool for the management of the pandemic.

The present study analyzed the variability of the *TMPRSS2* gene in individuals from two Brazilian Amazonian populations (Native Americans and admixed individuals from the city of Belém). Additionally, these results were integrated with compiled data from public databases, in order to trace a profile of the worldwide distribution of *TMPRSS2* variants potentially associated with the prevalence of the disease and the severity of the clinical symptoms presented by COVID-19 patients.

## 2. Results

The analyses of the exomes found in Amazonian populations revealed the presence of 41 variants of the *TMPRSS2* gene, including nine in codifying regions (two missense and seven synonyms) and 32 in non-codifying regions. The frequencies of these variants in the different continental populations are shown in Table 1.

**Table 1.** Description of the allele frequencies and consequence type recorded for the *TMPRSS2* in the both populations from north Brazil (NAM and BAP), the individuals from southeastern Brazil (ABraOM), and the continental populations from the 1,000 Genomes project (AFR, AMR, EAS, EUR, and SAS).

| Variant Id | Consequence | Minor Allele Frequency |  |  |  |  |  |  |  |
| --- | --- | --- | --- | --- | --- | --- | --- | --- | --- |
|  | Type | NAM | BAP | ABraOM | AFR | AMR | EAS | EUR | SAS |
| rs112132031 | Intron | 0.242 | 0.743 | 0.879 | 0.825 | 0.754 | 0.651 | 0.979 | 0.922 |
| rs140530035 | Intron | 0.250 | 0.730 | 0.883 | 0.768 | 0.746 | 0.655 | 0.977 | 0.917 |
| rs386416 | Intron | 0.226 | 0.581 | 0.662 | 0.568 | 0.561 | 0.302 | 0.700 | 0.646 |
| rs422471 | Intron | 0.226 | 0.562 | 0.660 | 0.568 | 0.561 | 0.301 | 0.698 | 0.647 |
| rs17854725 | Synonymous | 0.224 | 0.473 | 0.479 | 0.339 | 0.411 | 0.121 | 0.542 | 0.444 |
| rs2298660 | Intron | 0.155 | 0.317 | 0.215 | 0.387 | 0.223 | 0.250 | 0.201 | 0.179 |
| rs9974933 | Intron | 0.129 | 0.304 | 0.285 | 0.266 | 0.275 | 0.257 | 0.254 | 0.259 |
| rs9975014 | Intron | 0.129 | 0.304 | 0.285 | 0.266 | 0.275 | 0.257 | 0.254 | 0.263 |
| rs2298661 | Intron | 0.133 | 0.289 | 0.190 | 0.235 | 0.225 | 0.395 | 0.230 | 0.251 |
| rs2298659 | Synonymous | 0.172 | 0.267 | 0.185 | 0.175 | 0.218 | 0.249 | 0.230 | 0.189 |
| rs75603675 | Missense | 0.121 | 0.260 | 0.369 | 0.295 | 0.272 | 0.017 | 0.405 | 0.223 |
| rs55964536 | Intron | 0.017 | 0.239 | 0.361 | 0.101 | 0.288 | 0.006 | 0.483 | 0.398 |
| rs3787950 | Synonymous | NA | 0.169 | 0.126 | 0.218 | 0.061 | 0.155 | 0.079 | 0.257 |
| rs8126497 | Intron | 0.048 | 0.159 | 0.165 | 0.015 | 0.186 | 0.042 | 0.199 | 0.118 |
| rs12329760 | Missense | NA | 0.149 | 0.195 | 0.287 | 0.154 | 0.362 | 0.236 | 0.226 |
| rs73372193 | Intron | 0.016 | 0.054 | 0.039 | 0.197 | 0.024 | 0.001 | 0.002 | 0.001 |
| rs3819138 | Intron | NA | 0.047 | 0.083 | 0.009 | 0.088 | 0.049 | 0.151 | 0.071 |
| rs149695119 | 3'UTR | NA | 0.034 | 0.031 | 0.164 | 0.022 | NA | 0.003 | 0.002 |
| rs61735794 | Synonymous | NA | 0.020 | 0.022 | NA | 0.012 | 0.014 | 0.030 | 0.005 |
| rs73230068 | Intron | NA | 0.020 | 0.040 | 0.002 | 0.022 | NA | 0.039 | 0.006 |
| rs141788162 | Synonymous | NA | 0.014 | 0.001 | 0.002 | 0.006 | NA | 0.003 | NA |
| rs61735789 | Synonymous | NA | 0.014 | 0.014 | NA | 0.009 | NA | 0.013 | NA |
| rs188514624 | Intron | NA | 0.014 | 0.004 | 0.015 | 0.001 | NA | NA | NA |
| rs74423429 | Intron | NA | 0.014 | 0.012 | 0.001 | 0.007 | NA | 0.021 | 0.004 |
| * | Intron | NA | 0.007 | NA | NA | NA | NA | NA | NA |
| * | Intron | NA | 0.007 | NA | NA | NA | NA | NA | NA |
| * | Synonymous | NA | 0.007 | NA | NA | NA | NA | NA | NA |
| * | Intron | NA | 0.007 | NA | NA | NA | NA | NA | NA |
| * | Intron | NA | 0.007 | NA | NA | NA | NA | NA | NA |
| rs61735792 | Synonymous | NA | 0.007 | 0.013 | NA | 0.007 | NA | 0.017 | 0.002 |
| * | Intron | NA | 0.007 | NA | NA | NA | NA | NA | NA |
| rs141230106 | 3'UTR | NA | 0.007 | NA | NA | NA | 0.005 | NA | 0.001 |
| rs143712818 | Intron | NA | 0.007 | 0.002 | 0.015 | 0.001 | NA | NA | NA |
| rs75168613 | Intron | NA | 0.007 | 0.025 | 0.132 | 0.022 | 0.001 | 0.005 | 0.023 |
| rs113288437 | Intron | NA | 0.007 | 0.027 | 0.146 | 0.022 | 0.001 | 0.005 | 0.005 |
| * | Intron | NA | 0.007 | NA | NA | NA | NA | NA | NA |
| rs113928389 | Intron | NA | 0.007 | 0.002 | 0.015 | 0.001 | NA | NA | NA |
| rs75756279 | Intron | NA | 0.007 | 0.002 | NA | 0.001 | NA | 0.008 | 0.009 |
| rs144948620 | Intron | NA | 0.007 | 0.006 | NA | 0.001 | NA | 0.019 | 0.002 |
| * | Intron | NA | 0.007 | NA | NA | NA | NA | NA | NA |
| rs140141551 | Intron | 0.032 | NA | 0.015 | 0.001 | 0.010 | NA | 0.008 | 0.001 |

\*Id variant not characterized in literature; NA: Not Available; variant is absent in the referred population.

One of the missense mutations (rs12329760) is not found in the (NAM) investigated in the present study, while its frequency in the BAP population (0.149) is similar to that of the AMR population (0.159), but lower than that BAP in other populations, such as that of ABraOM (0.195). The other missense mutation (rs75603675) is present in both Amazonian samples, but has a reduced frequency in the NAM population (0.121) in comparison with the admixed population (0.26), as well as the population of ABraOM (0.37), although in all cases, the value was much higher than that recorded for the EAS population (0.017).

All the codifying variants of the 1,000 Genomes Project database and their respective allelic frequencies in the different continental populations were also evaluated. These variants included 388 missense, 26 stop-gain, 23 frameshift, 19 splice donor, nine splice acceptor, two inframe insertion, and two inframe deletion mutations (Detailed in Table S1).

Despite the considerable number of identified mutations most variants either occur at low frequencies or are absent from most of the populations evaluated. In fact, only two missense mutations, rs12329760 (0.261) and rs75603675 (0.244) have a Minor Allele Frequency (MAF) of over 0.01 in all the continental populations.

### 2.1. Variants that increase the expression of TMPRSS2 in the lungs

The distribution of the variations that potentially affect the expression of this gene in human tissue was also investigated. As infection with SARS-CoV-2 appears to begin with the upper respiratory tract, this investigation focused on the variants that alter substantially the expression of this gene in the pulmonary tissue.

The distribution of the eQTLs in the *TMPRSS2* gene was investigated based on the data of the Genotype-Tissue Expression Project (GTEx, https://www.gtexportal.org/home/datasets). A surprisingly large number (136) of mutations have a significant impact on the expression of *TMPRSS2* in the lungs. These mutations are listed in Table S2, which also shows the distribution of the allelic frequencies of the mutations in the populations from the different continents. The 39 markers that alter the expression of the TMPRSS gene in the lungs most significantly (p < 10-8) were deeply analyzed. Given the physical proximity of the markers, the occurrence of Linkage Disequilibrium (LD) was evaluated and blocks of variants were formed when they reached a high LD (D’ > 0.9; r2 > 0.8).

The application of this criterion resulted in the formation of seven blocks (I–VII) of variants that increase substantially the expression of *TMPRSS2* in the lungs. These data are presented in Table S3, which also shows the frequencies of these haplotypes in the populations from AFR, AMR, EAS, EUR, and SAS. All the frequencies were calculated from the data available in the 1,000 Genomes Project database.

Linkage Disequilibrium block I is formed by the following variants (with the allele responsible for the increase in the expression of *TMPRSS2* in the lungs): rs6517666 (allele A), rs6517668 (G), rs13050773 (A), rs1050080 (A), rs3737399 (T), rs3737400 (G), rs3737401 (T), rs13047535 (T), and rs13050726 (A). This block includes 12 haplotypes, although nine have a low frequency (<1%). By contrast, two haplotypes were responsible for 98% of the variability found in all the continental populations. The frequency of the Mutant Haplotype (MH) that contains all the mutations that increase the expression of *TMPRSS2* in the lungs is highest in AFR (0.606) and lowest in SAS (0.259). The EUR, AMR, and EAS populations have frequencies of 0.55-0.57, with no significant differences between them.

Linkage Disequilibrium block II includes five markers (and the alleles implicated in the increase in gene expression): rs363976 (allele G), rs468444 (A), rs469126 (T), rs469304 (A), and rs467798 (T). This block has 18 haplotypes, although, once again, most (15) have a reduced frequency (<1%). The haplotype that contains all the mutations that increase the expression of the gene (MH) is most frequent in the populations from EUR (53%), SAS (36%), and AMR (30%). The MH is very rare from EAS (<1%), however, and occurs at a low frequency in AFR (8%).

Linkage Disequilibrium block III is formed by six markers: rs467519 (allele A), rs467512 (G), rs468397 (C), rs401498 (C), rs35074065 (DEL) and rs463727 (A). Eleven haplotypes were identified, of which, eight have extremely low frequencies. Three haplotypes are responsible for more than 95% of the variability found among the different continents. In this block, Europe has the greatest variability, with four common haplotypes, and the highest frequency (0.425) of the MH. The frequency of the set of haplotypes that increase the expression of the gene is highest in EUR (0.565), followed by SAS (0.3864), AMR (0.316), and very much lower in AFR (0.124) and, in particular, EAS (0.007).

Linkage Disequilibrium block IV has three variants, rs2070788 (allele G), rs9974589 (A), and rs7364083 (G). Thus block has seven haplotypes, of which, three are practically restricted to the South Asian population. Three haplotypes represent 98% of all the continental variability, with one (IV.3) that is practically restricted to Africa. The haplotypes that contain variants that increase the gene expression are more frequent in the AMR (0.500), SAS (0.467), EUR (0.466) populations, and least frequent in the EAS (0.357) and AFR (0.293) populations.

Linkage Disequilibrium block V is formed by five markers: rs34624090 (allele INS), rs467375 (A), rs458213 (A), rs55964536 (T), and rs4818239 (C). This block has 14 haplotypes, of which, 10 are extremely limited. The European population has the greatest variability, with five common haplotypes, and the highest frequency (0.405) of the MH. The frequencies of the haplotypes that increase the expression of the gene are highest in EUR (0.540), followed by SAS (0.460), AMR (0.347), and AFR (0.255), with a minimal frequency in EAS (0.008).

Linkage Disequilibrium block VI has six markers: rs734056 (allele A), rs4290734 (G), rs139374762 (DEL), rs34783969 (T), rs11702475 (T), rs62217531 (T). This block has 21 haplotypes, although 13 have extremely low frequencies (<1%). The greatest variability is found in the African population, with 16 haplotypes, albeit the highest frequency recorded is 0.184 for the haplotype VI.1. The European populations nevertheless have the highest frequency of the MH haplotype (0.472), followed by the SAS (0.382), and AMR (0.288) populations. The East Asian populations share less than 1% of the haplotypes that increase the expression of the *TMPRSS2* gene.

Linkage Disequilibrium block VII is formed by five markers, rs383510 (allele T), rs35899679 (A), rs417888 (A), rs35041537 (T), and rs430915 (A). This group has 12 haplotypes, of which, eight have a reduced frequency. The EUR (0.460) and SAS (0.381) populations have the highest frequencies of the MH. The set of haplotypes that contain markers which increase gene expression had the highest frequency in the AMR (0.550) and EUR (0.501) populations, while the lowest frequencies were recorded in the EAS (0.430) and AFR (0.340) populations.

The proportions of the haplotypes which carry mutations that increase the expression of *TMPRSS2* in the lungs in the different populations are shown in Table 2. For each LD block, we computed the sum of frequencies of mutant haplotypes. In addition, the expression of *TMPRSS2* was computed concerning the mean of median of normalised expression from eQTL results in GTEx portal. Clearly, the European populations have the highest mean proportion of haplotypes that increase the expression of the gene (53%), followed by the populations from South Asia (46%) and America (42%). The lowest mean proportions of these haplotypes were recorded in Africa (28%) and, in particular, East Asia (19%). Specially, the alleles related with the greatest expression of *TMPRSS2* in lung (24 of the 39 markers, p < 10-8) have only a residual frequency (< 1%) in the East Asians (Table S2).

**Table 2.** Haplotypes block frequency and *TMPRSS2* expression. For each LD block, the sum of frequencies of mutant haplotypes and the expression of *TMPRSS2* was computed concerning the mean of the median of normalized expression from eQTL results in GTEx portal.

| Haplotype | Expression | AFR | AMR | EAS | EUR | SAS |
| --- | --- | --- | --- | --- | --- | --- |
| Blocks | value | Frequency | Frequency | Frequency | Frequency | Frequency |
| I | 0.210 | 0.2753 | 0.5591 | 0.5119 | 0.5755 | 0.6196 |
| II | 0.228 | 0.2776 | 0.3213 | 0.0069 | 0.5378 | 0.3701 |
| III | 0.241 | 0.1241 | 0.3156 | 0.0069 | 0.5656 | 0.3865 |
| IV | 0.164 | 0.2927 | 0.4986 | 0.3571 | 0.4662 | 0.4673 |
| V | 0.195 | 0.2549 | 0.3473 | 0.0079 | 0.5398 | 0.4591 |
| VI | 0.189 | 0.4206 | 0.3343 | 0.0099 | 0.495 | 0.4417 |
| VII | 0.140 | 0.3404 | 0.5461 | 0.4286 | 0.507 | 0.4652 |
| **<u>X</u>** |  | **0.2837** | **0.4175** | **0.1899** | **0.5267** | **0.4585** |

In addition, the different haplotypes groups (I–VII) were examined to determine whether they varied in their levels of expression of *TMPRSS2* in the lungs (Table 2). The data indicate that Linkage Disequilibrium (LD) block III has the set of markers that most express the *TMPRSS2* gene in the lungs, with a mean of 0.241. The second highest level of expression was recorded in LD block II (mean = 0.228). Pairwise analysis of *TMPRSS2* expression were performed between each pair of haplotype by ANOVA. The expression in the II and III is significantly higher than those of the others (p-value = 8.86e-10).

## 3. Discussion

Over the past 20 years, beta-coronaviruses have caused severe global epidemics, including those provoked by SARS-CoV in 2002, MERS-CoV in 2012, and the current outbreak (COVID-19), caused by SARS-CoV-2, which began in 2019 and became a pandemic in 2020 [1, 2]. The epidemiological data available on COVID-19 indicate considerable variation in susceptibility to the disease, clinical outcomes, and mortality rates, which is most apparent in the comparison between the continents of Europe, East Asia, and Africa [9]. Within Europe, in addition, major differences are found among countries. While it is possible that the incidence is under-reported in some countries, the epidemiological data indicate clearly that the genetic variability of different human populations may contribute to the differences in both the incidence of the disease and its mortality rates.

The genetic predisposition of humans for most disorders, including infectious diseases, such as those caused by viruses, usually involves a complex of genes, each of which may contribute, even in a minor way, to the occurrence or severity of the disease [28, 29].

The *ACE2* and *TMPRSS2* genes appear to be two likely components of the potential genetic predisposition of humans to COVID-19, given that the combined expression of these two genes is necessary to allow the virus to access human cells [18, 19]. A number of studies of the association between the variability of the *ACE2* gene and both susceptibility to infection and the clinical outcome of COVID-19 patients are currently under way [16, 20, 21]. However, no variants of this gene appear to be capable of altering the expression of this gene in the lungs (GTEx) [30].

The mutations of the *TMPRSS2* gene that may be associated with the outcome of COVID-19 may result from (i) functional mutations, which provoke a qualitative shift in the capacity of the virus to access the human cell, and (ii) quantitative modifications of gene expression, which modify the number of potential sites available for the virus to access the interior of the cells.

Both potential mechanisms were investigated in detail here, including the analysis of the two samples from Brazilian Amazonia, the Native American population and the admixed population, through the complete sequencing of the exomes, together with the analysis of two databases, the ABraOM database [31], which contains data on 609 individuals from southeastern Brazil, and the 1,000 Genomes Project [32], which includes 2,504 individuals from 26 populations on five continents.

The inclusion of data on Native American populations in the present study is fundamentally important, considering the known susceptibility of these populations to external pathogens, as well as the limited data available on the genetic variability of these populations. The results of the analysis revealed reduced variability in the Amazonian populations in terms of the functional modifications of the *TMPRSS2* gene, which included only two missense mutations that had alternative allele frequencies of over 1%. This reduced variability is a common feature of the populations of all the continents, however, and despite the long list of functional mutations that were identified, only two – the same ones identified in the Native American populations – had frequencies of the alternative allele greater than 1%. These findings indicate that functional mutations likely have only a negligible impact on the frequency or severity of COVID-19.

The analyses of the genetic alterations that modify the expression of *TMPRSS2* in human pulmonary cells revealed variants with a potential clinical role [33, 17]. The initial analyses revealed 136 genetic markers that alter gene expression significantly, including 39 with high levels of differential expression among the genotypes (p < 10-8). The markers that presented significant Linkage Disequilibrium (D’ > 0.90 and r2 > 0.8) were assigned to one of seven blocks (I–VII) and analyzed according to their haplotypes.

The analysis of the expression-modifying variants of the *TMPRSS2* gene in the Amerindians (NAM) and the admixed population from Brazilian Amazon (BAP) revealed the presence of only four (rs17854725, rs55964536, rs9974933, and rs9975014) of the 136 genetic markers that alter the pulmonary expression of *TMPRSS2*. A possible explanation for the reduced number of such variants in both populations is the data extrapolation from exome analyses, which is unable to cover all the genome regions. Due to the paucity of the data, this study focuses on the analysis of expression variants in the populations of 1,000 Genomes.

Considerable levels of variation were observed among the populations. The analyses of all the Linkage Disequilibrium (LD) blocks indicated that the European populations have the highest proportion of haplotypes that increase the expression of the *TMPRSS2* gene, conversely, the lowest proportions of the haplotypes that increase gene expression were recorded in East Asia.

The combined analysis of the 39 genetic markers investigated here revealed that the populations from Europe and South Asia may have, on average, greater pulmonary expression of the *TMPRSS2* gene than the other continental populations. This implies that these populations would be the most vulnerable to COVID-19 infection, based on this parameter.

By contrast, the evidence presented here indicates that the pulmonary expression of *TMPRSS2* may be more reduced among individuals from the East Asian populations. This is based on the much lower mean frequencies of the high-expression haplotypes in the different blocks and, in particular, the residual frequencies of these haplotypes (< 1%) in the two blocks with the greatest additive effect, in the EAS. These populations would thus have a reduced risk of viral infection.

## 4. Materials and Methods

### 4.1. Study population

The study population is composed of 74 Native Americans and 94 admixed individuals from the Amazon region of northern Brazil. The Native Americans represent 10 different Amazonian ethnic groups, which were grouped together as the Native American (NAM) group. The 94 admixed individuals live in Belém, a city located in northern Brazil, and were grouped together as the Brazilian Admixed Population (BAP). Rodrigues et al., (2020) provides further details on these populations [34].

We compared our results with those of the populations included in the phase 3 release of the 1,000 Genomes Project (available at http://www.1000genomes.org) [32]. These data included populations of African (AFR), Admixed American (AMR), East Asian (EAS), European (EUR), and South Asian (SAS) descent. Data on the genomic variants in Brazilian populations were also obtained from the Online Archive of Brazilian Mutations (ABraOM), freely available at http://abraom.ib.usp.br [31].

### 4.2. Ethics committee approvals

The present study was approved by the Brazilian National Committee for Ethics in Research (CONEP) and the Research Ethics Committee of the UFPA Tropical Medicine Center, under CAAE number 20654313.6.0000.5172. All the participants, together with the tribal leaders, signed a free informed-consent form, and whenever necessary, a translator explained the project and its importance to the participants. The participants were recruited between September 2017 and December 2018.

### 4.3. Extraction of the DNA and preparation of the exome libraries

The genetic material was extracted using the phenol-chloroform method described by Sambrook et al. 1989 [35]. The quantity of the DNA was assessed using a Nanodrop-8000 spectrophotometer (Thermo Fisher Scientific Inc., Wilmington, DE, USA) and its integrity was determined by electrophoresis in 2% agarose gel.

The Nextera Rapid Capture Exome (Illumina) and SureSelect Human All Exon V6 (Agilent) kits were used to prepare the exome libraries, following the manufacturer’s instructions. The NextSeq 500® platform (Illumina®, US) along with the NextSeq 500 High-output v2 300 cycle kit (Illumina®) were used to run the sequence reactions.

### 4.4. Bioinformatic analysis

The quality of the FASTQ reads was analyzed (FastQC v.0.11-http://www.bioinformatics.babraham.ac.uk/projects/fastqc/), and the samples were filtered to eliminate low-quality readings (fastx_tools v.0.13 - http://hannonlab.cshl.edu/fastx_toolkit/). The sequences were mapped and aligned with the reference genome (GRCh37) using the BWA v.0.7 tool (http://bio-bwa.sourceforge.net/), and the file was then indexed and sorted (SAMtools v.1.2 - http://sourceforge.net/projects/samtools/). This alignment was processed for the removal of duplicate PCRs (Picard Tools v.1.129 - http://broadinstitute.github.io/picard/), mapping quality recalibration, and local realignment (GATK v.3.2 - https://www.broadinstitute.org/gatk/). These results were processed to distinguish the variants from the reference genome (GATK v.3.2).

### 4.5. Statistical analyses

The means and p-values of the expression levels of the polymorphisms of the *TMPRSS2* gene were obtained by the Genotype-Tissue Expression (GTEx) project (available at: https://www.gtexportal.org) [30]. The allele frequencies of the NAM and BAP populations were obtained directly by gene counting and were compared with those of the other study populations (AFR, EUR, AMR, EAS, SAS, and ABraOM). The Linkage Disequilibrium (LD) was estimated and the haplotype blocks were compiled using the “LDlink” web tool [36], accessed at http://analysistools.nci.nih.gov/LDlink/. The differences in gene expression among the haplotype blocks (I-VII) were evaluated by a one-way Analysis of Variance (ANOVA), and the significance of the difference between the most divergent blocks was determined by Tukey’s HSD test. All analyses were run in RStudio v.3.5.1, using the “multcompView” package, v.0.1-8 [37].

## 5. Conclusions

The presented data indicates a potential genetic contribution to the susceptibility of the individual to viral infection and might also influence the outcome of COVID-19, based on genotypic variations which influence the expression of *TMPRSS2* in the lungs. These findings are in accordance with the epidemiological data, which report a higher incidence and mortality rates in Europeans, who have a greater constitutive expression of the gene, while a lower incidence and mortality rates are found in populations with a lower constitutive expression of the gene, in particular, from East Asia.

Further research will be needed to validate the possible associations identified here, and the potential clinical applications of this information. The identification of specific groups, or even individuals, at greater risk of developing COVID-19 would support the development of specific public health strategies and initiatives appropriate for combating COVID-19.

## Data Availability

- The 1,000 Genomes Project (available at http://www.1000genomes.org)
- Online Archive of Brazilian Mutations (ABraOM)
(available at http://abraom.ib.usp.br).

http://www.1000genomes.org

http://abraom.ib.usp.br

## Supplementary Materials

Supplemental Table.

## Author Contributions

Conceptualization, N.P.C.S. and S.E.B.S.; methodology, A.M.R.S., A.R.S. and J.C.G.R.; formal analysis, P.P., L.F.P., F.C.M. and G.S.A.; investigation, J.A.G.M., M.R.F., B.C.M.K. and P.B.A.; writing—original draft preparation, N.P.C.S., A.S.K., J.C.G.R., A.R.-Dos-S. and S.E.B.S ; writing—review and editing, N.P.C.S., J.C.G.R, A.S.K., A.M.R.S., P.P.A, G.S.A, A.R.-Dos-S. and S.E.B.S; supervision, N.P.C.S., A.S.K., P.P.A. and S.E.B.S. All authors have read and agreed to the published version of the manuscript.

